# Early-life inflammatory markers and subsequent episodes of depression and psychotic experiences in the ALSPAC birth cohort

**DOI:** 10.1101/2022.07.12.22277542

**Authors:** A. J. Edmondson-Stait, X. Shen, M. J. Adams, M. C. Barbu, H. J. Jones, V. E. Miron, J. Allardyce, J. P. Boardman, S. M. Lawrie, A. M. McIntosh, G. M. Khandaker, A. S. F. Kwong, H. C. Whalley

## Abstract

**Background:** Inflammation is implicated in depression and psychosis, including association of childhood inflammatory markers on the subsequent risk of developing symptoms. However, it is unknown whether early-life inflammatory markers are associated with the number of depressive and psychotic symptoms from childhood to adulthood.

**Methods:** Using the prospective Avon Longitudinal Study of Children and Parents birth cohort (N=up-to 6,401), we have examined longitudinal associations of early-life inflammation [exposures: interleukin-6 (IL-6), C-reactive protein (CRP) levels at age 9y; IL-6 and CRP DNA-methylation (DNAm) scores at birth and age 7y; and IL-6 and CRP polygenic risk scores (PRSs)] with the number of depressive episodes and psychotic experiences (PEs) between ages 10-28 years. Psychiatric outcomes were assessed using the Short Mood and Feelings Questionnaire and Psychotic Like Symptoms Questionnaires, respectively. Exposure-outcome associations were tested using negative binomial models, which were adjusted for metabolic and sociodemographic factors.

**Results:** Serum IL-6 levels at age 9y were associated with the total number of depressive episodes between 10-28y (n=4,262; β=0.086; 95%CI:0.036-0.137; p_FDR_=0.009). CRP DNAm score at birth was associated with total number of PEs, size but this association did not survive correction for multiple testing (n=822; β=0.204; 95%CI:0.024-0.388; p_uncorrected_=0.027; p_FDR_=0.252). Other immune measures were not associated with depression or PEs.

**Conclusions:** Early-life inflammatory markers are associated with the burden of depressive episodes and of PEs subsequently from childhood to adulthood. These findings support a potential role of early-life inflammation in the aetiology of depression and psychosis and highlight inflammation as a potential target for treatment and prevention.

## Introduction

Peripheral low-grade chronic inflammation, measured by the levels of cytokines and acute phase proteins such as interleukin-6 (IL-6) and C-reactive protein (CRP) in the serum, have been cross-sectionally associated with many psychiatric disorders, including major depressive disorder and schizophrenia (1). However, the role of these inflammatory biomarkers in the aetiology of these disorders remains inconclusive. It is hypothesised that inflammation may be on causal pathways to depression and psychosis. Mendelian randomisation (MR) studies, that assess causality using genetic instruments, have indeed provided evidence for inflammation being on the causal pathway to psychiatric disorders, rather than the reverse (2).

Longitudinal studies have also demonstrated that higher levels of serum IL-6 and CRP in childhood are associated with increased risk of psychotic disorders and depression in early-adulthood, consistent with MR studies (2-6). Previous studies have shown associations between inflammation and subsequent persistent depressive symptoms. Investigating such severity and burden is important since multiple depressive episodes have been shown to associate with a more severe depressive phenotype and treatment resistance (7, 8) and persistent PEs are associated with developing severe mental health problems(9). Here, we extend these studies by investigating whether early-life inflammation associates with the burden of depressive episodes and PEs, measured during a longer follow-up period up to 28 years of age.

Additionally, we use multiple ways of indexing early-life inflammation. Previous studies have typically assessed only serum measures of CRP and IL-6 to investigate the associations between inflammation and psychiatric disorders. However, these serum measures can fluctuate and are affected by factors such as BMI, recent infections, medication and other inflammatory conditions(10, 11). Genetic and epigenetic predictors of immune proteins have been shown to be more robust for assessment of such factors, and provide a more stable/long-term proxy for the proteins they predict, compared to serum measures (12, 13). DNA methylation (DNAm) scores and polygenic risk scores (PRSs) can be used as indicators of an individual’s epigenetic and genetic risk respectively to a trait or phenotype, or in this case protein level. We use multiple measures of CRP and IL-6, by not only assessing protein levels in serum, but also investigating DNAm scores and PRSs of CRP and IL-6 from multiple early-life time points, to robustly assess the effect of these proteins.

In this study we aimed to determine whether inflammation earlier in life can predict the total number of depressive episodes and PEs observed from ages 10 to 28 years. We utilised a longitudinal cohort, Avon Longitudinal Study of Children and Parents (ALSPAC), with biomarkers of inflammation measured in childhood and prospective data on depressive episodes and PEs measured at 16 time points throughout adolescence into early adulthood (ages 10-28 years). We hypothesised that both acute (serum) and longer-term (DNAm scores and PRSs) biomarkers of inflammation will associate with multiple depressive episodes and PEs.

## Methods and Materials

### Study Sample

Pregnant women resident in Avon, UK with expected dates of delivery 1st April 1991 to 31st December 1992 were invited to take part in The Avon Longitudinal Study of Children and Parents (ALSPAC) (14-16). The total sample size for analyses using any data collected after the age of seven is therefore 15,454 pregnancies, resulting in 15,589 foetuses. Of these 14,901 were alive at 1 year of age. Further details are described in Supplementary Methods. Demographics of sample individuals used within the current study are shown in tables 1 to 3.

**Table 1.** Demographics of participants with inflammatory blood marker data. Includes participants with missing data.

| Variable | Female | Male |
| --- | --- | --- |
|  | N = 2,477 | N = 2,522 |
| Maternal Education |  |  |
| CSE/O-level/Vocational | 1,167<br>(47%) | 1,239<br>(49%) |
| A-level/Degree | 1,012<br>(41%) | 1,022<br>(41%) |
| Missing | 298 (12%) | 261 (10%) |
| BMI (age 9 years) | 17.80<br>(2.91) | 17.36<br>(2.60) |
| Missing | 35 | 19 |
| CRP (age 9 years) | 0.66 (1.08) | 0.46 (0.95) |
| IL-6 (age 9 years) | 1.35 (1.50) | 1.11 (1.39) |
| Total Depressive Episodes |  |  |
| 0 | 1,169<br>(47%) | 1,720<br>(68%) |
| 1 | 504 (20%) | 403 (16%) |
| 2 | 283 (11%) | 158 (6.3%) |
| 3 | 178 (7.2%) | 71 (2.8%) |
| 4 | 125 (5.0%) | 30 (1.2%) |
| 5 | 77 (3.1%) | 20 (0.8%) |
| 6 | 42 (1.7%) | 9 (0.4%) |
| 7 | 21 (0.8%) | <5 (0.2%) |
| 8 | 11 (0.4%) | <5 (<0.1%) |
| 9 | 8 (0.3%) | <5 (0%) |
| 10 | <5 (<0.1%) | <5 (0%) |
| Missing | 58 (2.3%) | 106 (4.2%) |
| Total PEs |  |  |
| 0 | 1,987<br>(80%) | 2,076<br>(82%) |
| 1 | 284 (11%) | 200 (7.9%) |
| 2 | 77 (3.1%) | 47 (1.9%) |
| 3 | 26 (1.0%) | 9 (0.4%) |
| 4 | 7 (0.3%) | <5 (0.2%) |
| 5 | 9 (0.4%) | <5 (0%) |
| 6 | <5 (0.1%) | <5 (<0.1%) |
| 7 | <5 (<0.1%) | <5 (0%) |
| Missing | 82 (3.3%) | 185 (7.3%) |
| n (%); Mean (SD) |  |  |

**Table 2.** Demographics of participants with inflammatory DNAm score data. Includes number of participants with missing data for each variable.

| Variable | Female | Male |
| --- | --- | --- |
|  | N = 514 | N = 484 |
| Maternal Education |  |  |
| CSE/O-level/Vocational | 242 (47%) | 221 (46%) |
| A-level/Degree | 245 (48%) | 245 (51%) |
| Missing | 27 (5.3%) | 18 (3.7%) |
| BMI (age 7 years) | 16.31 (2.30) | 16.16 (1.82) |
| Missing | <5 | <5 |
| CRP DNAm score (at birth) | -0.026<br>(0.008) | -0.028<br>(0.008) |
| Missing | 49 | 44 |
| CRP DNAm score (age 7 years) | -0.018<br>(0.007) | -0.019<br>(0.008) |
| Missing | 24 | <5 |
| IL-6 DNAm score (at birth) | -7.12 (0.72) | -7.26 (0.69) |
| Missing | 49 | 44 |
| IL-6 DNAm score (age 7 years) | -7.20 (0.71) | -7.20 (0.69) |
| Missing | 24 | <5 |
| Total Depressive Episodes |  |  |
| 0 | 222 (43%) | 280 (58%) |
| 1 | 107 (21%) | 101 (21%) |
| 2 | 71 (14%) | 54 (11%) |
| 3 | 52 (10%) | 19 (3.9%) |
| 4 | 23 (4.5%) | 13 (2.7%) |
| 5 | 15 (2.9%) | 10 (2.1%) |
| 6 | 14 (2.7%) | <5 (0.2%) |
| 7 | 5 (1.0%) | <5 (0.2%) |
| 8 | <5 (0.4%) | <5 (0%) |
| 10 | <5 (0.2%) | <5 (0%) |
| Missing | <5 (0.4%) | 5 (1.0%) |
| Total PEs |  |  |
| 0 | 419 (82%) | 407 (84%) |
| 1 | 66 (13%) | 59 (12%) |
| 2 | 16 (3.1%) | 13 (2.7%) |
| 3 | <5 (0.4%) | <5 (0.2%) |
| 4 | <5 (0.8%) | <5 (0%) |
| 5 | <5 (0.8%) | <5 (0%) |
| 6 | <5 (0.4%) | <5 (0%) |
| Missing | <5 (0.2%) | <5 (0.8%) |
| n (%); Mean (SD) |  |  |

**Table 3.** Demographics of participants with inflammatory PRSs data. Includes number of participants with missing data for each variable.

| Variable | Female | Male |
| --- | --- | --- |
|  | N = 3,825 | N = 4,022 |
| Maternal Education |  |  |
| CSE/O-level/Vocational | 1,854 (48%) | 2,002 (50%) |
| A-level/Degree | 1,433 (37%) | 1,449 (36%) |
| Missing | 538 (14%) | 571 (14%) |
| CRP PRS | 0.0000<br>(0.0000) | 0.0000<br>(0.0000) |
| IL-6 PRS | 0.0000<br>(0.0000) | 0.0000<br>(0.0000) |
| Total Depressive Episodes |  |  |
| 0 | 1,610 (42%) | 2,189 (54%) |
| 1 | 683 (18%) | 534 (13%) |
| 2 | 345 (9.0%) | 184 (4.6%) |
| 3 | 252 (6.6%) | 97 (2.4%) |
| 4 | 165 (4.3%) | 43 (1.1%) |
| 5 | 98 (2.6%) | 32 (0.8%) |
| 6 | 59 (1.5%) | 11 (0.3%) |
| 7 | 32 (0.8%) | 6 (0.1%) |
| 8 | 20 (0.5%) | <5 (0%) |
| 9 | 8 (0.2%) | <5 (<0.1%) |
| 10 | <5 (<0.1%) | <5 (0%) |
| Missing | 550 (14%) | 925 (23%) |
| Total PEs |  |  |
| 0 | 2,738 (72%) | 2,738 (68%) |
| 1 | 395 (10%) | 270 (6.7%) |
| 2 | 109 (2.8%) | 64 (1.6%) |
| 3 | 37 (1.0%) | 16 (0.4%) |
| 4 | 8 (0.2%) | <5 (<0.1%) |
| 5 | 12 (0.3%) | <5 (<0.1%) |
| 6 | 5 (0.1%) | <5 (<0.1%) |
| 7 | <5 (<0.1%) | <5 (0%) |
| Missing | 519 (14%) | 927 (23%) |
| n (%); Mean (SD) |  |  |

#### Psychiatric Outcomes

##### Measures of depressive episodes

The Short Mood and Feelings Questionnaire (SMFQ) was used to assess self-reported depressive symptoms at 11 time points between the ages of 10 to 28 years (Supplementary Figures 1-2, Supplementary Table 1). The SMFQ was administered via the mail or in clinics. There were three clinic time points (ages 10, 12 and 14 years) and eight remote self-reported (mail) time points (ages 17, 18, 19, 22, 23, 24, 26 and 28 years). The SMFQ measures depressive symptoms experienced in the past 2 weeks and comprises of 13 questions. Each question response is a from 0 to 2, where the total summed score ranges from 0 to 26. A depressive episode is defined as a score >= 11, as this cut off has previously been shown to have good specificity for predicting depression (17, 18). Total depressive episodes were calculated by summing the occurrence of a depressive episode at each time point.

### Measures of PEs

The Psychotic Like Symptoms Questionnaire (PLIKS-Q) was used to assess psychotic like experiences at 9 time points between the ages of 13 to 26 years of age (Supplementary Figures 1-2, Supplementary Table 2). PLIKS-Q was administered via the mail or in clinics. There were three clinic time points (ages 12, 18 and 24 years) and six remote self-reported (mail) time points (ages 11, 13, 14, 16, 21 and 26 years). PLIKS-Q asks about the presence, frequency and context of experiences associated with psychosis. A PE is defined as answering “Yes - Definitely” to questions asking if the participant had heard anything others had not or seen anything others had not, or if the participant had answered “Yes - Definitely” to questions asking if they felt they were be spied upon or followed and that this occurred at least once a month. This has previously been used as a measure of PEs in ALSPAC (19). Total number of PEs were then calculated by summing the occurrence of a PE at each time point.

Sensitivity analyses were conducted to test whether using an interviewer rated definition of PEs, available at the three clinic time points, gave consistent results to the primary analyses. Using additional PLIKS-Q questions asked during the clinic assessments, interviewers rated PE symptoms as either not present, suspected or definitely present. From this a binary variable was derived: definite PE or no/suspected PE. Total number of interviewer rated PEs were calculated for the three clinic time points and regression analysis was then conducted as in the main analysis.

Sensitivity analyses were conducted to test whether an interviewer rated definition of PEs, available at the three clinic time points, gave consistent results to the primary analyses. Using additional PLIKS-Q questions asked during the clinic assessments and these were used to define an alternative definition of PEs determined by the interviewer (binary variable: definite PE or no/suspected PLE). using this interviewer rated definition for PEs. Regression analysis was then conducted as in the main analysis.

### Inflammatory Exposures

#### Serum IL-6 and CRP

Blood samples were collected from individuals at age 9 years (N = 5,079; mean age: 9.86 years; SD: 0.31) and high sensitivity serum CRP and serum IL-6 were measured as described previously (3). CRP and IL-6 were found to be the most commonly increased inflammatory markers in a meta-analysis of inflammatory serum markers across psychiatric disorders (1). Consistent with previous studies(3), individuals with serum CRP >= 10mg/L were removed from further analysis (N = 60), as this level of CRP would indicate current acute inflammation. Serum CRP and IL-6 were log transformed to achieve a normal distribution of residuals.

#### IL-6 and CRP DNA methylation

DNA methylation was quantified in a subsample of individuals (N = 998) from blood samples obtained from cord blood at birth and at age 7 years (mean age: 7.45; SD: 0.13), as described previously (20, 21). DNA methylation scores were calculated by multiplying the DNA methylation M-value with the effect size for each CpG on a phenotype (obtained through independent association analyses), and then summed. Effect sizes for 7 and 35 CpGs, previously shown to associate with CRP and IL-6 respectively in independent samples, were used to calculate the respective DNAm scores (12, 22). The CRP DNAm score has previously been used and validated in ALSPAC (21). There was no overlap in participant samples used to estimate the effect sizes used to generate the CRP and IL-6 DNAm scores with ALSPAC. White blood cell (WBC) type proportion estimates (B-cells, CD4 T-cells, CD8 T-cells, granulocytes, monocytes and natural killer cells) were estimated from DNA methylation data using the Houseman method(23). This uses a prior reference data to estimate WBC proportions in whole blood samples. DNAm principal components (PCs) were calculated by first residualizing standardised DNA methylation M-values (after removing 44,171 cross-reactive or polymorphic probes(24)) on age, sex and array, and then applying principal component analysis (PCA) on these residuals.

#### IL-6 and CRP PRSs

Publicly available genome-wide association study (GWAS) summary statistics were downloaded to calculate PRSs. GWASs on circulating levels of CRP and IL-6 in the blood were respectively obtained from UK Biobank (N = 343,524) (downloaded from the Neale lab repository – http://www.nealelab.is/uk-biobank/) and Finish cohorts (total N = 8,233) (The Cardiovascular Risk in Young Finns Study and FINRISK) (25). Where available, GWASs underwent quality control by removing SNPs with minor allele frequency (MAF) < 0.01 and INFO (imputation quality) < 0.8. INFO was not supplied for IL-6 GWAS summary statistics. There was no sample overlap between either of the GWASs used and the ALSPAC cohort.

Genotyping information (including quality control procedures) of ALSPAC has been described in detail elsewhere and is detailed in the Supplementary Methods (26). PRSs were calculated using SbayesR (27) on unrelated participants (N = 7,975). SBayesR is a Bayesian method that adjusts the beta values in the GWAS based on LD scores from a reference panel. Shrunk sparse LD matrices based on 1.1 million common SNPs in a random sample of 50K unrelated European individuals were downloaded from the GCTB website (https://cnsgenomics.com/software/gctb/) and used as the reference panel. Default values were used for the variables “pi”, “gamma”, “chain-length”, “burn-in” and “out-freq”, (see code available on GitHub). Additional arguments were parsed to the function including “ambiguous-snp” (removes SNPs with ambiguous nucleotides, ie. A/T or G/C), “imputing N” (imputes per-SNP sample size) and “exclude-MHC” (excludes SNPs in the major histocompatibility complex (MHC regions) - Chr6:28-34 Mb). The number of SNPs used to calculate PRSs for CRP and IL-6 were 286,512 and 1,129,461 respectively.

### Statistical analysis

All continuous variables were standardised using z-score scaling to obtain standardised effect sizes (β). Negative binomial models were used to test the associations between inflammatory serum markers (age 9 years), DNAm scores (age 0 and 7 years) and PRSs with subsequent total number of depressive episodes (age 10 to 28 years) and PEs (age 13 to 26 years). Two main models were used, the first was a base model covarying for sex only and secondly a fully adjusted model which included BMI (for serum and DNAm scores at age 9 and 7 years respectively) and maternal education (Figure 1). For the DNAm analysis the base and fully adjusted models also included methodological covariates of 10 DNAm informed PCs and the fully adjusted model additionally included methodological covariates of DNAm WBC estimates. For the PRS analysis the base and fully adjusted models also included 10 genetically informed PCs to adjust for population stratification. Sex, maternal education and BMI were used as covariates as these have all been previously shown to be associated with inflammation or psychiatric symptoms (28, 29). Maternal education was coded as a binary variable as either “CSE/O-level/Vocational education” or “A-level/degree level of education”. Sex was coded as a binary variable as either “Male” or “Female”. BMI (age 7 and 9 years) was calculated by dividing weight (kg) by squared height (meters). Genetic principal components were calculated using PLINK (30).

**Figure 1.**
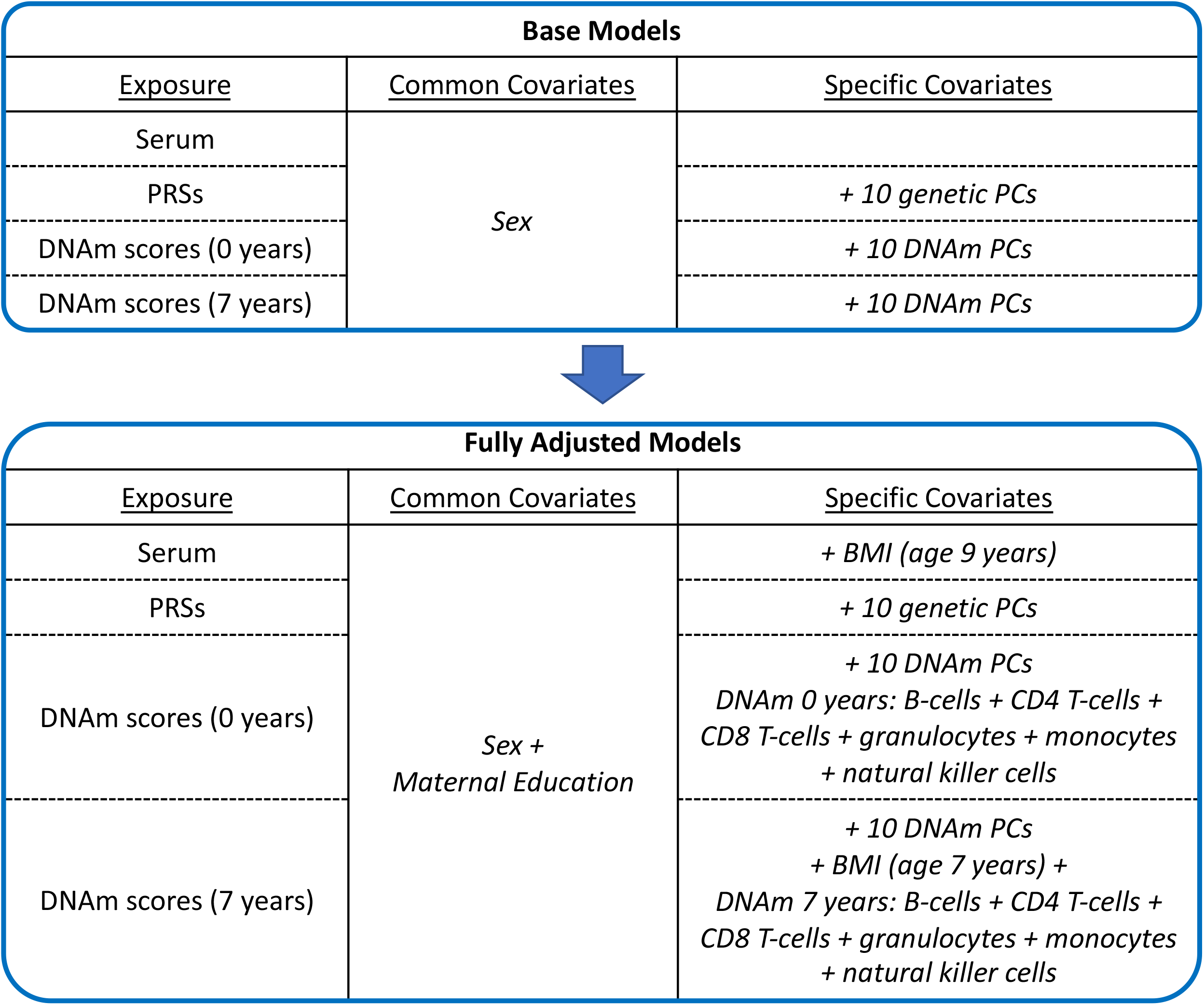
Covariates used in base and fully adjusted models. Common and specific covariates used for each different exposure in base and fully adjusted models.

P-values were corrected for multiple testing using the false discovery rate (FDR) method and significance was deemed FDR corrected p-value (p_FDR_) < 0.05. 95% CIs are reported throughout.

Code used for analysis is openly available at www.github.com/AmeliaES/ALSPAC_inflam_2022.

### Sensitivity analyses

Pearson’s correlation coefficients were calculated to test for correlations between serum CRP and IL-6 with DNAm scores and PRSs of CRP and IL-6.

Other sensitivity analyses included sex stratification analysis, as sex has been shown to be an important factor associated with inflammation in the context of psychiatric outcomes (28). In these models, sex was no longer included as a covariate.

### Imputation of missing outcome and covariate data

To address potential bias from sample attrition additional analysis was run using Multiple Imputation Chained Equations (MICE) to impute missing data for outcomes and covariates (this is detailed in Supplementary Methods)(31).

## Results

### Sample characteristics of total depressive episodes and PEs from childhood to adulthood

In the subsample of individuals with CRP or IL-6 serum data (total N = 5,069) 51% of females and 23% of males had experienced at least one depressive episode and 17% of females and 11% of males had experienced at least one PE over 13 years (Table 1). In the subsample of individuals with DNAm scores (total N = 998), 57% of females and 41% of males had experienced at least one depressive episode and 18% of females and 15% of males had experienced at least one PE (Table 2). In the subsample of individuals with PRSs (total N = 8,788), 43% of females and 23% of males had experienced at least one depressive episode and 15% of females and 9% of males had experienced at least one PE (Table 3). Similar percentages of individuals experienced depressive episodes or PEs across the samples used.

### Pearson’s correlation of inflammatory serum markers with PRS and DNAm scores

Serum IL-6 at age 9 years correlated with IL-6 PRS (r = 0.037; p = 0.014; N = 4536) and IL-6 DNAm scores at birth (r = 0.101; p = 0.009; N = 683) but not age 7 years (r = -0.004; p = 0.920; N = 732). Serum CRP at age 9 years correlated with CRP PRS (r = 0.149; p < 0.001; N = 4493), but not CRP DNAm scores at birth (r = 0.042; p = 0.275; N = 672) or age 7 years (r = 0.073; p = 0.050; N = 721). Serum inflammatory biomarkers associate with PRSs more than DNAm scores.

### Associations between inflammation measures and total depressive episodes and PEs

There was strong evidence that serum IL-6 was associated with the total number of depressive episodes (ages 10 to 28 years) in both the base (β=0.093; 95% CI:0.045-0.141; p_FDR_=0.001) and fully adjusted models (β=0.0864; 95% CI:0.0359-0.137; p_FDR_ = 0.006) (Figure 2A; Supplementary Table 3). However, we found little evidence for associations between other inflammatory biomarkers (serum CRP, DNAm scores and PRSs) and total depressive episodes (Figure 2A; Supplementary Table 3).

**Figure 2.**
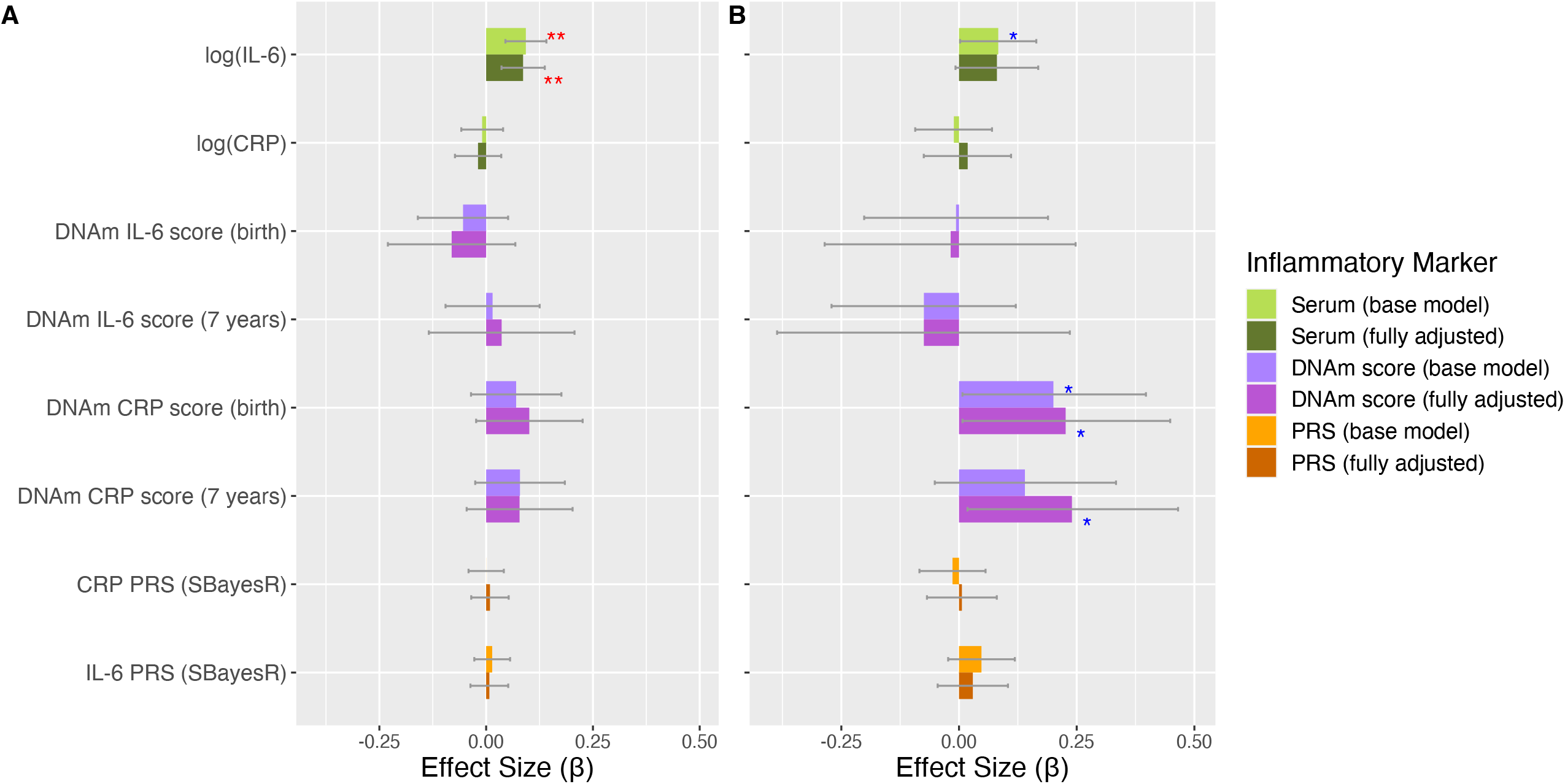
Association of inflammatory markers with total number of A) depressive episodes and B) PEs. Standardised effect sizes from negative binomial models with 95% CIs displayed as bars. Red asterisks indicate significance using FDR corrected p-values, blue asterisks indicate significance using uncorrected p-values. Levels of significance: *: p < 0.05; **: p < 0.01.

There was little evidence of association between inflammatory biomarkers and total number of PEs (ages 13 to 26 years) (Figure 2B; Supplementary Table 3). However, we observed weak associations (p_uncorrected_ < 0.05) between CRP DNAm scores derived from blood at birth and the total number of PEs in both the base model (β=0.201; 95% CI:0.007-0.397; p_uncorrected_=0.042; p_FDR_=0.167) and the fully adjusted model (β=0.226; 95% CI: 0.008-0.449; p_uncorrected_= 0.044; p_FDR_=0.176). Similar effect sizes were observed for CRP DNAm score derived from blood at birth in a sensitivity analysis using an alternative definition of PEs (interviewer rated using additional PLIKS-Q questions) available at three clinic assessments (ages 12, 18 and 24 years) (Supplementary Figure 4; Supplementary Table 5). Weaker associations were also observed for associations with serum IL-6 (base model: β=0.083; 95% CI: 0.002-0.164; p_uncorrected_=0.040; p_FDR_= 0.167) and DNAm scores derived from bood at age 7 years (fully adjusted model: β=0.240; 95% CI: 0.0181-0.465; p_uncorrected_=0.0364; p_FDR_=0.176), however this was only observed in either the base or fully adjusted model rather than both (Figure 2B; Supplementary Table 3). We found little evidence of association between the other inflammatory biomarkers and total number of PEs (Figure 2B; Supplementary Table 3).

Results of other associations are described in Supplementary Table 3. Similar effect sizes were observed when missing data was imputed (Supplementary Table 3; Supplementary Figure 3). Similar effect sizes were also observed between serum IL-6 and total depressive episodes in females and males for both base and fully adjusted models (Supplementary Table 3; Figure 3).

**Figure 3.**
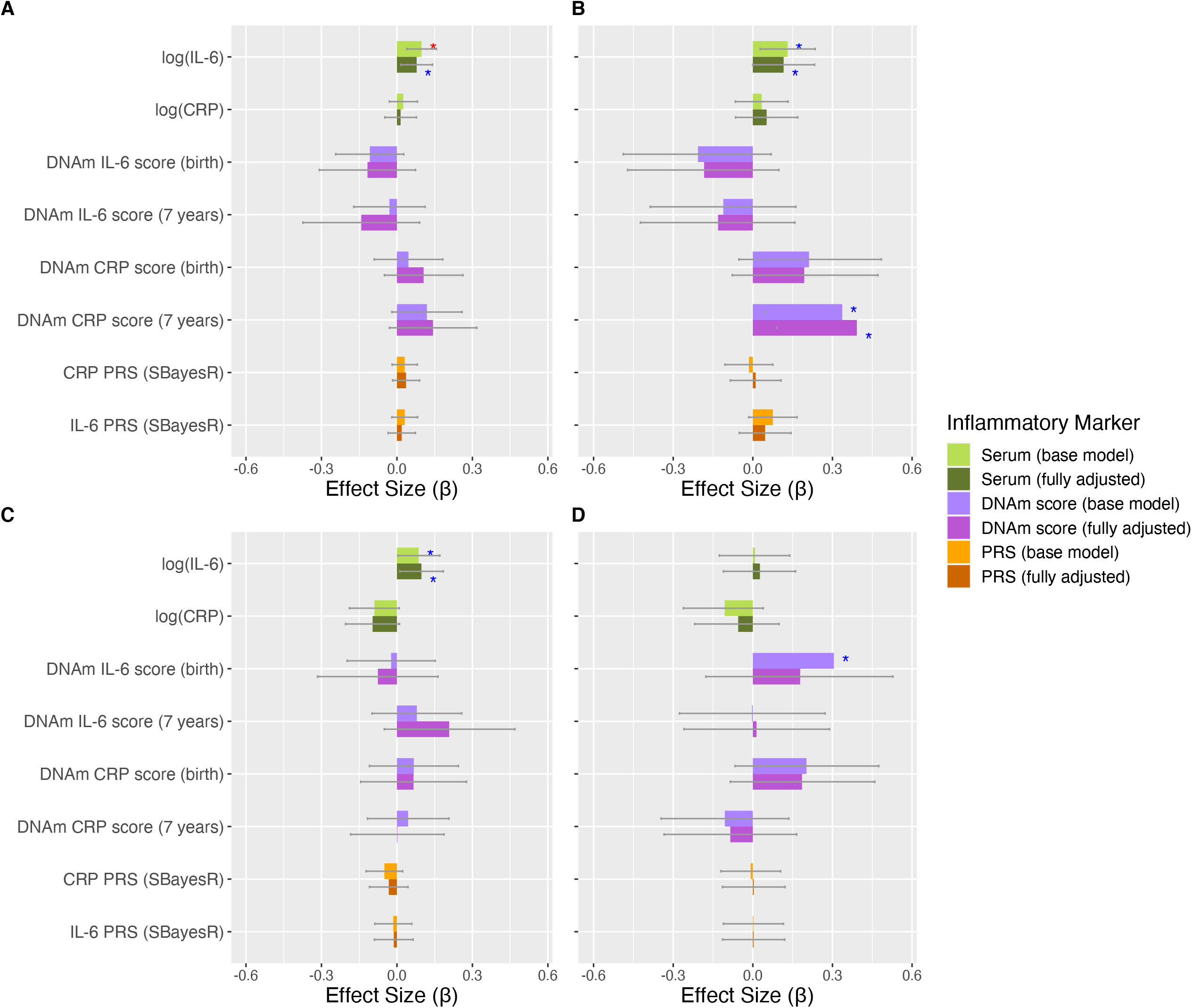
Association of inflammatory markers with total number of depressive episodes and PEs, split by sex. In females, A) total number of depressive episodes and B) PEs as outcomes. In males, C) total number of depressive episodes and D) PEs as outcomes. Standardised effect sizes from negative binomial models with 95% CIs displayed as bars. Red asterisks indicate significance using FDR corrected p-values, blue asterisks indicate significance using uncorrected p-values. Level of significance: *: p < 0.05.

## Discussion

To our knowledge, this is one of the first studies to investigate early-life inflammation (acute and stable biomarkers) with the subsequent burden of depression and PEs, across early adolescence and into the third decade of life. This follow-up period includes the peak incidence period for major psychiatric disorders and continued neurodevelopment. We found childhood inflammation, as measured by serum IL-6 at age 9 years, was associated with the subsequent burden of depressive episodes (up to 28 years). Further, biomarkers of inflammation at birth, as measured by CRP DNAm, were weakly related to the burden of PEs later in life (up to 26 years).

Our findings are consistent with previous studies in ALSPAC (3),(4). For instance, serum IL-6 at age 9 years has been shown to associate with depression and PEs at age 18 years (3) and depressive episodes, psychotic disorder and negative symptoms at age 24 years (4). Our results extend these findings, by using multiple timepoints of mental health assessments (N=16) to assess the effect of early-life inflammation on the burden of psychiatric outcomes. Additionally, we utilised novel biomarkers of inflammation that represent a more stable/long-term measure of inflammation. We show that childhood inflammation is associated with the total number of depressive and psychotic experiences from childhood to adulthood. Multiple episodes of depression are an important marker of disorder burden and severity as they are associated with later chronic depression and treatment resistance (7, 8). Persistent PEs, as opposed to ones that are transient, are also associated with a greater general psychopathology and increased risk for psychotic disorders (9, 32, 33). Hence it is important to investigate in detail over multiple time points what precedes and predicts frequent occurrences of depression and PEs across this critical period of development of these disorders, to better understand risk factors for burden and severity of illness.

Similar to the above studies in ALSPAC (3),(4), we did not find associations between the acute inflammatory biomarker serum CRP and later life psychiatric outcomes, potentially due to measurement fluctuations of serum CRP. Many previous studies investigating the impact of inflammation on psychiatric outcomes only include acute serum biomarkers of inflammation. However, these biomarkers fluctuate throughout the day and are heavily influenced by recent infection (34). We therefore investigated novel epigenetic and genetic biomarkers of inflammation, which reflect a longer-term, more stable inflammatory exposure. Other studies, including our own investigating adult population cohorts, have highlighted the importance of researching DNAm markers of inflammation over serum-based markers (12, 35, 36). CRP DNAm scores associated with more brain regions and with larger effect sizes than serum CRP, in the context of depression (36). Further, CRP DNAm scores were also better predictors of cognition than serum measures of CRP (35). Here, we found weak associations between CRP DNAm scores derived at birth and total number of PEs. These associations had larger effect sizes (β) than associations with serum measures but were conducted in smaller samples, which may have contributed to the wide confidence intervals observed and thus lack of significance after correcting for multiple tests. CRP DNAm scores derived from blood at age 7 years had smaller effect sizes than scores derived at birth and showed no evidence for associations in the base model. However, when additionally adjusting for BMI, maternal education and WBC estimates these effect sizes increased, possibly due to multicollinearity. Stronger associations observed for DNAm scores derived from birth rather than age 7 years may be because DNA methylation across the genome is highly sensitive to experiences around the time of birth and this is associated with changes in brain connectivity (37). Additionally, systemic inflammation in the new-born period is strongly associated with atypical brain development (38, 39). Our findings extend the current literature providing preliminary evidence that stable/long-term biomarkers of CRP could be important indicators for the burden of mental health outcomes, though further studies in larger samples would be required to confirm this relationship. Further studies should also investigate how early life DNAm markers of inflammation affect brain development, which may contribute to the development of psychiatric illness.

We observed strong associations between serum IL-6 and total number of depressive episodes, and weaker associations with total number of PEs. MR studies have indicated IL-6 to be potentially causal for schizophrenia and depression (2). Additionally, a recent MR study suggests that IL-6 may have causal effects on brain structures relevant to psychiatric disorders (40). Therefore, inflammation early in childhood may be associated with depressive episodes and PEs through mediating changes in brain structure and function. Future studies should extend this work further to investigate this. Despite similar effect sizes being observed for both associations of serum IL-6 with total number of depressive episodes and PEs, only the associations with total depressive episodes remained after correcting for multiple testing and fully adjusting for potential confounders. This could be due to differences in power to detect associations with depressive episodes compared to PEs. Depressive episodes were more common than PEs in our sample which would increase power for detecting associations between inflammation and depressive episodes (Supplementary Figure 2). It is also for this reason that we investigated the total number of PEs across several assessments rather than an outcome of persistent PEs across all time points, despite persistent PEs perhaps being more clinically meaningful (33). A lack of relationship between inflammatory biomarkers and total number of PEs could also relate to methodological issues, such as the difference in definitions for identifying PEs between studies. Previous ALSPAC studies have shown associations between inflammation and subsequent PEs at single time points at age 18 and 24 years (3, 4). These used an interviewer-determined definition of PEs, based on additional PLIKS-Q questions, available at clinic appointments. Our study maximises data availability by including responses from six remote assessments in addition to the three clinic appointments. For consistency, we only used PLIKS-Q questions available at all time points to define PEs, similar to a previous ALSPAC study(19). We observed similar effect sizes between the main analysis and a sensitivity analysis using the interviewer-rated definition of PEs at the three clinic time points only (Supplementary Figure 4, Supplementary Table 5). This suggests our findings were not influenced by using different PE definitions.

Despite inflammatory PRSs correlating with their serum equivalents, we did not find associations between inflammatory PRSs and depressive episodes or PEs. This is similar to another study in a large cohort of young individuals (41). This could be due to potential weak power of the PRSs, due to small discovery sample sizes for the GWASs. Additionally, ancestry is important to take into acount and is a possible source of bias for PRS analysis. The IL-6 PRS may have been particularly prone to this as the IL-6 GWAS was conduced in Finnish cohorts, which may have different ancestry structure to participants in ALSPAC. Further, PRSs scores were projected from association studies conducted in older individuals. It is known that immune system function changes with age (42). This may also explain the weaker associations observed with DNAm scores. Therefore, although we validated our PRSs and DNAm scores by testing for correlations with their serum equivalents, this difference in ages between the discovery samples and ALSPAC cohort may also have influenced the associations observed between PRSs and DNAm with psychiatric outcomes. Future studies should also conduct GWASs and epigenome wide association studies (EWASs) in younger cohorts so scores can be calculated from cohorts of similar ages. Finally, using a larger selection of inflammatory biomarkers, rather than being limited to only CRP and IL-6, will also help in understanding which inflammatory pathways are important in the context of psychiatric disorders.

Potential bias from sample attrition is a potential threat for all longitudinal studies, causing missing data (43). Therefore, we imputed missing data for sex, maternal education, BMI (age 7 and 9 years) and depression SMFQ scores and PEs at each time point using multiple imputation (31). We observed similar effect sizes in the imputed datasets analysis to the complete case main analysis, indicating our results are robust to this potential source of bias.

Our study used the same measures of depression and PEs from 16 assessment points across 18 years, making it one of the most detailed longitudinal studies in a large sample. We were able to go beyond simple acute serum-based biomarkers of inflammation and expand to additionally exploring PRSs and DNAm scores. This enabled us to further our understanding of the effect of inflammation on disorder burden within this important developmental period in adolescence to early adulthood. Our work builds upon the existing evidence showing inflammation in early childhood to be important for psychiatric burden later in life.

## Supporting information

Supplementary Infomation

## Data Availability

The ALSPAC study website contains details of all data available through a fully searchable data dictionary (http://www.bristol.ac.uk/alspac/researchers/our-data/).

## Acknowledgements

This research was funded in whole, or in part, by the Wellcome Trust (Grant No. 108890/Z/15/Z). For the purpose of open access, the authors have applied a CC BY public copyright licence to any Author Accepted Manuscript version arising from this submission.

The UK Medical Research Council and Wellcome (Grant No.: 217065/Z/19/Z) and the University of Bristol provide core support for ALSPAC. This publication is the work of the authors and Amelia Edmondson-Stait and Alex Kwong will serve as guarantors for the contents of this paper. A comprehensive list of grants funding is available on the ALSPAC website (http://www.bristol.ac.uk/alspac/external/documents/grant-acknowledgements.pdf); This research was specifically funded by the MRC (MR/M006727/1 and GO701503/85179), Wellcome Trust (08426812/Z/07/Z)], Wellcome Trust and MRC (092731), NIH (PD301198-SC101645).

We are extremely grateful to all the families who took part in this study, the midwives for their help in recruiting them, and the whole ALSPAC team, which includes interviewers, computer and laboratory technicians, clerical workers, research scientists, volunteers, managers, receptionists and nurses.

GWAS data was generated by Sample Logistics and Genotyping Facilities at Wellcome Sanger Institute and LabCorp (Laboratory Corporation of America) using support from 23andMe.

MCB is supported by a Guarantors of Brain Non-clinical Post-Doctoral Fellowship. HJJ is supported by the NIHR Biomedical Research Centre at University Hospitals Bristol and Weston NHS Foundation Trust and the University of Bristol. The views expressed are those of the authors and not necessarily those of the NIHR or the Department of Health and Social Care. AMM acknowledges funding support from the Wellcome Trust Innovator Award (Grant No. 220857/Z/20/Z). GMK acknowledges funding support from the Wellcome Trust (Grant No. 201486/Z/16/Z), the Medical Research Council (Grant No. MC_PC_17213, Grant No. MR/S037675/1, and Grant No. MR/W014416/1), The MQ: Transforming Mental Health (Grant No. MQDS17/40), and the BMA Foundation (J. Moulton Grant 2019).

