## Supplementary Infomation for "Early-life inflammatory markers and subsequent episodes of depression and psychotic experiences in the ALSPAC birth cohort"

Supplementary Methods

**ALSPAC Cohort**

Study data (1-3) were collected and managed using REDCap electronic data capture tools hosted at the University of Bristol (4). REDCap (Research Electronic Data Capture) is a secure, web-based software platform designed to support data capture for research studies. Please note that the study website contains details of all the data that is available through a fully searchable data dictionary and variable search tool" and reference the following webpage: <http://www.bristol.ac.uk/alspac/researchers/our>[-data/](http://www.bristol.ac.uk/alspac/researchers/our-data/). Ethical approval for the study was obtained from the ALSPAC Ethics and Law Committee and the Local Research Ethics Committees. Consent for biological samples has been collected in accordance with the Human Tissue Act (2004). Informed consent for the use of data collected via questionnaires and clinics was obtained from participants following the recommendations of the ALSPAC Ethics and Law Committee at the time.

**Quality control and processing of genetic data**

Genetic data was processed by ALSPAC according to standard procedures which are as follows: ALSPAC children were genotyped using the Illumina HumanHap550 quad chip genotyping platforms by 23andme subcontracting the Wellcome Trust Sanger Institute, Cambridge, UK and the Laboratory Corporation of America, Burlington, NC, US. The resulting raw genome-wide data were subjected to standard quality control methods. Individuals were excluded on the basis of gender mismatches; minimal or excessive heterozygosity; disproportionate levels of individual missingness (>3%) and insufficient sample replication (IBD < 0.8). Population stratification was assessed by multidimensional scaling analysis and compared with Hapmap II (release 22) European descent (CEU), Han Chinese, Japanese and Yoruba reference populations; all individuals with non-European ancestry were removed. SNPs with a minor allele frequency of < 1%, a call rate of < 95% or evidence for violations of Hardy-Weinberg equilibrium (P < 5E-7) were removed. Cryptic relatedness was measured as proportion of identity by descent (IBD > 0.1). Related subjects that passed all other quality control thresholds were retained during subsequent phasing and imputation. 9,115 subjects and 500,527 SNPs passed these quality control filters.

**Multiple Imputation of missing covariate and outcome data**

Multiple imputation chained equations was conducted using the "mice" package in R (5) to impute missing data for sex, maternal education, BMI (age 7 and 9 years) and depression SMFQ scores and PEs at each appointment time point. Three imputations were conducted for serum, DNAm scores and PRSs samples separately where the samples were subsetted to complete respective outcome data. All variables in the substantive model were included in the multiple imputation. Additional auxiliary variables were included in the multiple imputation if they predicted the variable to impute or it’s missingness (to reduce the bias of variables being "missing not at random") or if they had < 40% missing data. N auxiliary variables included: BMI (age 12 years), DV: SDQ emotional symptoms score (prorated) (age 7, 9, 11 and 13 years), DV: Yes-no anxiety disorder (parent computer prediction, ICD-10 and DSM-IV) (age 10 and 13 years), DV: DAWBA DSM-IV clinical diagnosis - Any anxiety disorder (age 7 years), DV: 13yr Yes-no depressive disorder (parent computer prediction, ICD-10 and DSM-IV), WEMWBS Composite score (age 23 years), IMD Score 2010, quintiles; at timepoint Jan 2014 (YP) and maternal Edinburgh Post-Natal Depression score. One hundred imputed datasets were created. Total depressive episodes and PEs were then calculated in each imputed dataset to be used in the substantive model. Effect sizes from each imputed dataset were then pooled using Rubin’s rule. Sample sizes following imputation compared against sample sizes used in the complete case analysis are shown in Supplementary Table 4.

Supplementary Figures

**Supplementary Figure 1. Mean ages of individuals at each appointment time point.** Mean age plotted as a point on violin plots of age distributions.

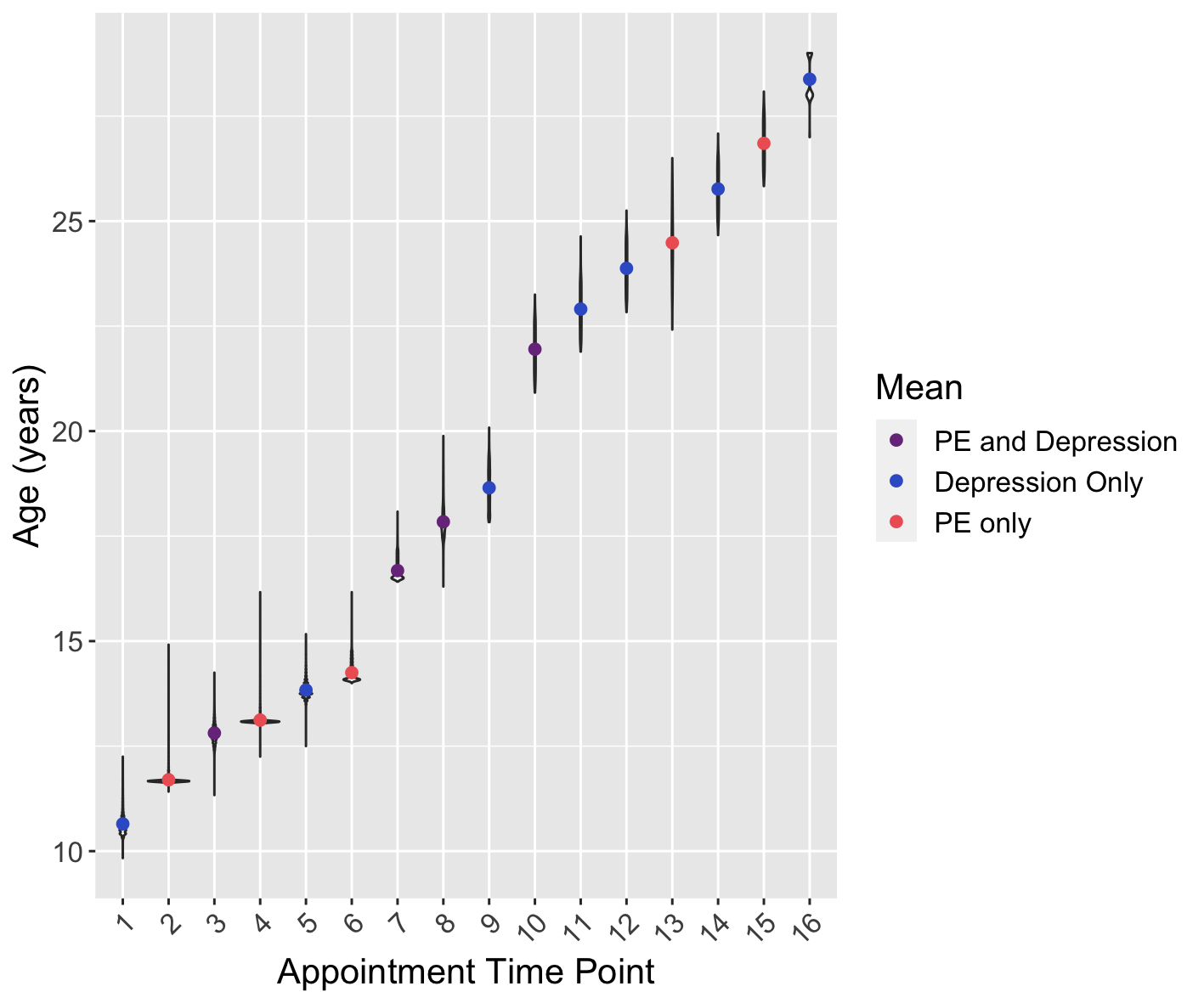

**Supplementary Figure 2. Number of participants with depressive episodes and PEs.** Number of participants at each appointment timepoint (same time points as Supplementary Figure 1) that have experienced A) a depressive episode and B) a PLE. Total number of participants and the total number of C) depressive episodes and D) PEs they have experienced over all appointment time points.

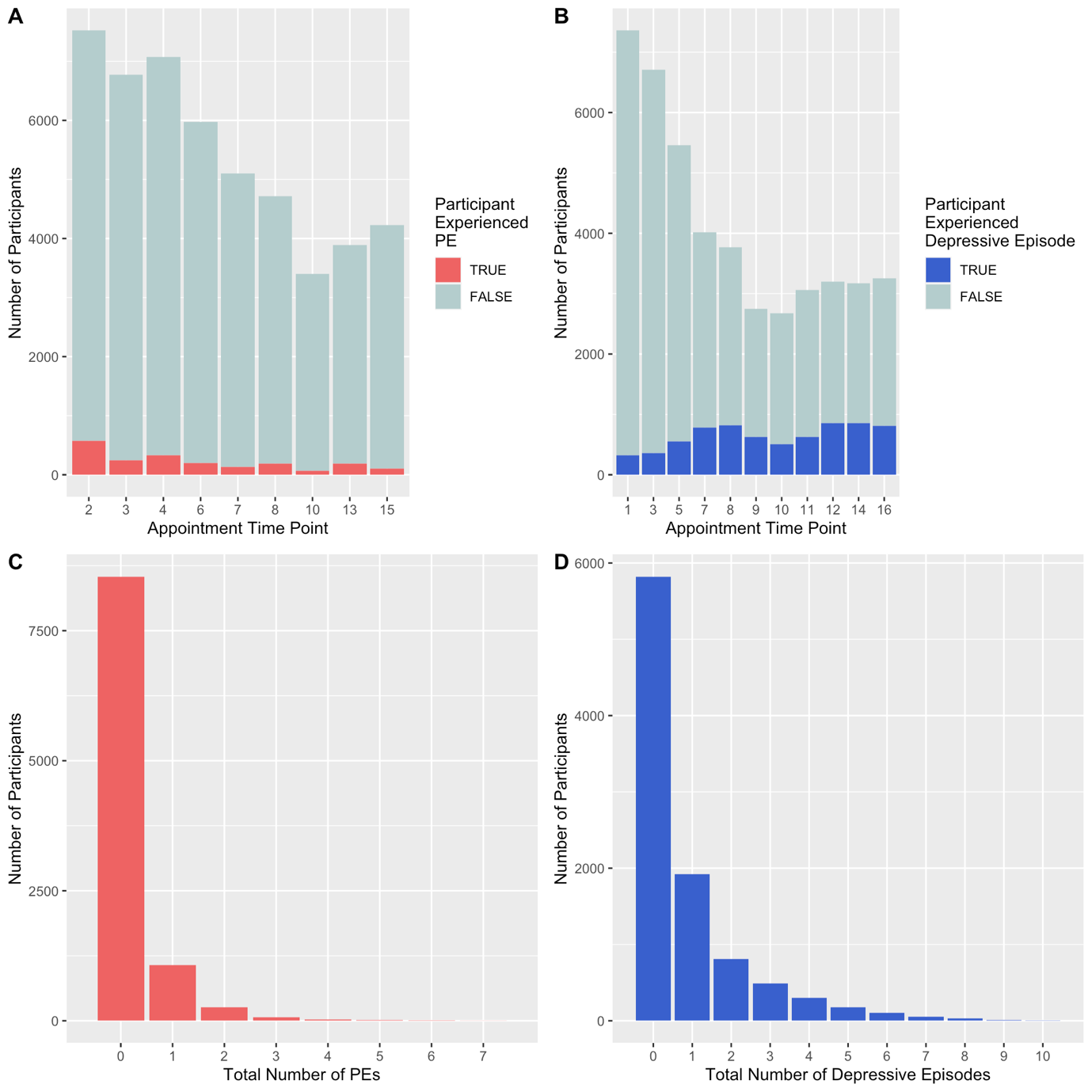

**Supplementary Figure 3. Association of inflammatory markers with total number of A) depressive episodes and B) PEs on imputed dataset.** Standardised effect sizes from negative binomial models with 95% CIs displayed as bars. Red asterisks indicate significance using FDR corrected p-values, blue asterisks indicate significance using uncorrected p-values. Levels of significance: *: p < 0.05; **: p < 0.01.

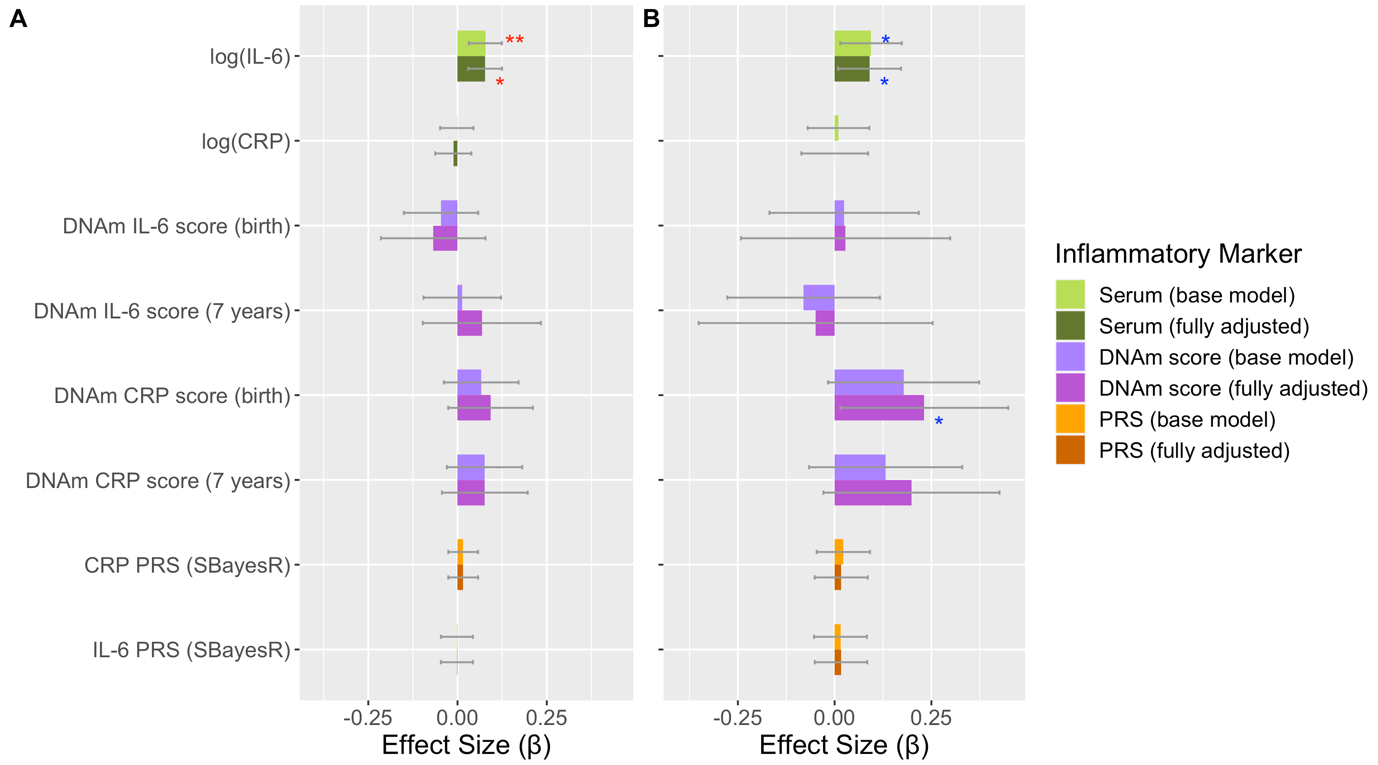

**Supplementary Figure 4. Associations of inflammatory markers with total number of PEs at clinic time points assessed at ages 12, 18 and 24 years.** PEs defined from PLIKS-Q interview interviewer rated “definite” PLE. Bars represent 95% confidence intervals. Blue asterisks indicate significance using uncorrected p-values. Level of significance: *: p < 0.05

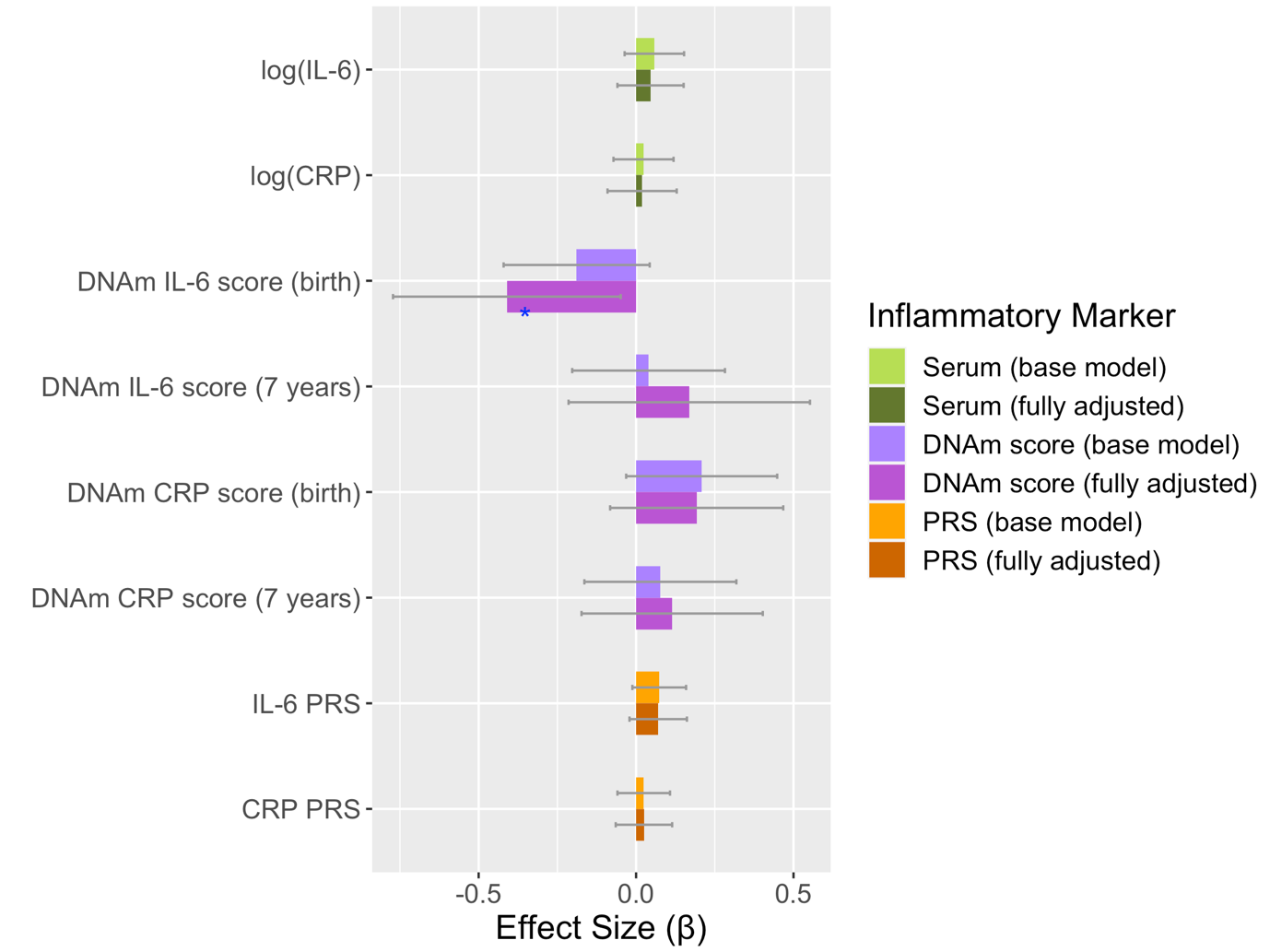

Supplementary Tables

**Supplementary Table 1. Descriptive statistics of SMFQ responses in total sample.** Columns refer to occasion corresponds to appointment time point shown in Figures 1 and 2. Sample size, mean, standard deviation (SD), median, interquartile range (IQR) and percentage of those with a score above or equal to the thresholds core of 11.

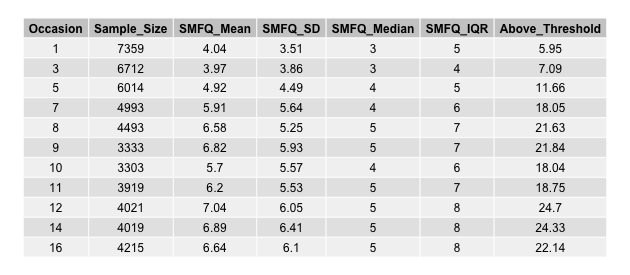

**Supplementary Table 2. Descriptive statistics of presence of PE in total sample.** Occasion corresponds to appointment time point shown in Figures 1 and 2. Columns refer to sample size, whether participant experienced a PE at that time point, or no PLE, percentage of individuals that experienced a PLE, number of participants with missing data on all questions used to determine whether participant experienced a PE and whether the responses were acquired by a clinic appointment or were self-reported.

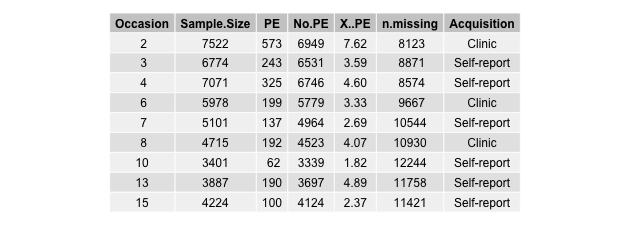

**Supplementary Table 3. Association of inflammatory markers with total number of depressive episodes and PEs.** Includes base and fully adjusted models, split-by-sex analysis and multiple imputation results.

*See Supp_Tab3.xlsx*

**Supplementary Table 4. Sample sizes for models.** Including sample sizes from main analysis (base and fully adjusted) compared with those used for the multiple imputation models where missing data was imputed.

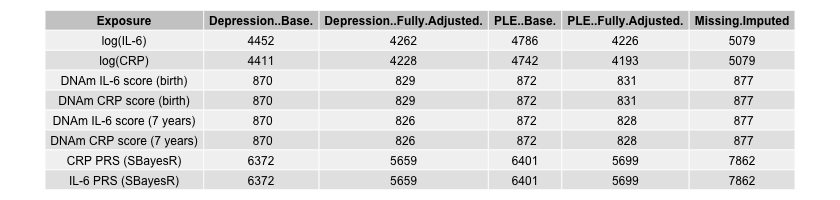

**Supplementary Table 5. Associations of inflammatory markers with total number of PEs at clinic time points (ages 12, 18 and 24 years).** Including PEs defined from PLIKS-Q interviewer rated definition of having a “definite” PLE.

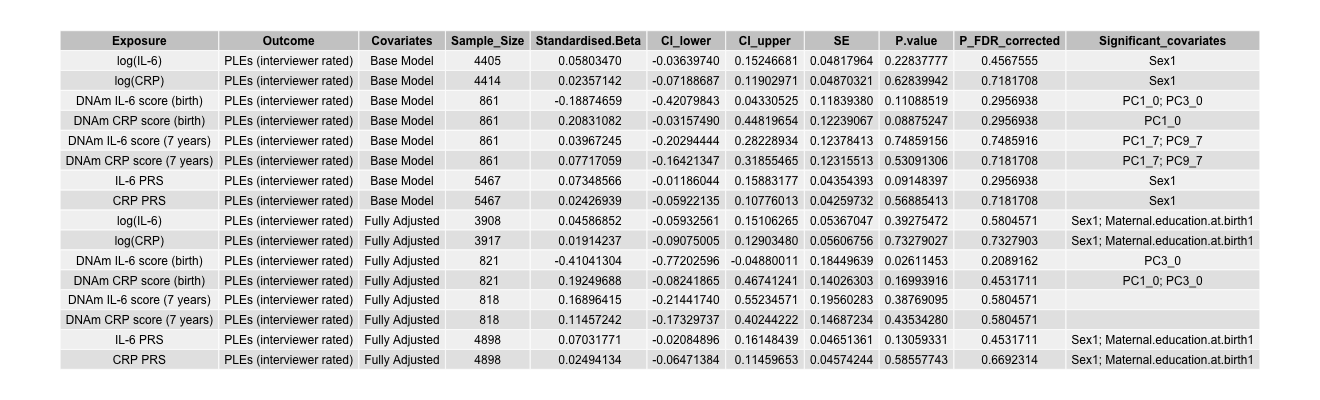
